# Anti-Interleukin-23 Treatment Linked to Improved *Clostridioides difficile* Infection Survival

**DOI:** 10.1101/2024.12.03.24318323

**Authors:** Gregory R. Madden, Robert Preissner, Saskia Preissner, William A. Petri

## Abstract

*Clostridioides difficile* is a leading cause of healthcare associated infection and an unacceptably high proportion of patients with *C. difficile* infection die despite conventional antibiotic treatment. Host-directed immunotherapy has been proposed as an ideal treatment modality for *C. difficile* infection to mitigate the underlying toxin-mediated pathogenic immune response while sparing protective gut microbes. Interleukin-23 monoclonal antibody inhibitors are used extensively to control pro-inflammatory Th17 immune pathways in psoriasis and inflammatory bowel disease that are similarly important during *C. difficile* infection. We used a large retrospective electronic health record database to test the hypothesis that hospitalized patients with *C. difficile* infection who are on anti-IL-23 treatment will have improved survival compared to patients without anti-IL-23. 9,301 anti-IL-23 patients had significantly lower probability of all-cause death within 30 days (0.54%) compared with 1:1 propensity-matched control patients (3.1%). IL-23 inhibition is a promising adjunct to *C. difficile* treatment and further clinical trials repositioning anti-IL-23 monoclonal antibodies from psoriasis and inflammatory bowel disease to *C. difficile* infection are warranted.

## Introduction

*Clostridioides difficile* is one of the leading healthcare associated pathogens associated with unexpected in-hospital death.^1^ *C. difficile* produces a diarrheal toxin which stimulates a strong pathogenic immune response, resulting in death in 5% or more of cases.^2^ Recent advances in microbiota and anti-toxin therapies given after recovery from *C. difficile* infection are proven to restore a healthy gut microbiome and prevent recurrent infection but do not address early severe outcomes, including death.

Antibiotics in *C. difficile* infection (including those used against *C. difficile*) are a double-edged sword because they disrupt the protective gut microbiota and predispose to further *C. difficile* acquisition, growth, and reinfection. Clinical options for augmenting treatment in severe or refractory *C. difficile* infection remain extremely limited and each lack strong evidence to support their efficacy: 1) changing or combining anti-*C. difficile* antibiotics, 2) changing antibiotic administration route (per rectum, via ileostomy), 3) early fecal microbiota transplant (rarely done during acute infection due to limited evidence and logistical barriers) and/or 4) intestinal surgery.^3^

IL-23 signaling induces colitis in mouse models of inflammatory bowel disease^4^ and, similarly, interleukin-23 (IL-23)-mediated Th17 immunity plays a pivotal role in neutrophil activation and severity of *C. difficile* infection.^5^ IL-23-positive immune cells infiltrate the human colon during *C. difficile* colitis^6^ and higher IL-23 correlates with inflammasome activation^7^ and worsening disease severity.^8^ IL-23 neutralization in mice prevents *C. difficile* infection mortality^6^ and IL-23 inhibitors have been proposed as a promising candidate for host-directed immunotherapy in *C. difficile* infection.^9^

Beginning with ustekinumab (FDA approved in 2009), monoclonal antibodies against IL- 23 (specifically, the pro-inflammatory p19 subunit, IL-23p19) have become a mainstay of treatment for psoriasis and inflammatory bowel disease. Here we report a retrospective, propensity-matched case control study of *C. difficile* infection associated 30-day mortality in patients receiving anti-IL-23 treatment compared to patients without anti-IL-23.

## Method

Data were collected from the COVID-19 Research Network provided by TriNetX which includes 94 healthcare organizations in 11 countries. The time window used for this analysis was between 20 years and 30 days prior to the query on October 9, 2024. Hospitalized *C. difficile* infection cases were defined as inpatient encounters with a positive *C. difficile* test (polymerase chain reaction or enzyme immunoassay) and/or International Classification of Diseases-10 (ICD-10) code A04.72 (‘Enterocolitis due to *Clostridium difficile*, not specified as recurrent’). Anti-IL-23 therapy was defined as receipt of any available anti-IL-23 treatment (ustekinumab, guselkumab, tildrakizumab, risankizumab, and mirikizumab).

Baseline demographic, comorbidity (based on ICD-10 codes) and laboratory data were collected and two-tailed t-tests were used to determine the statistical differences between groups. Outcomes analyses were performed after 1:1 propensity score matching for age and sex. The primary outcome was all-cause death (as recorded in the medical record system from contributing sites) within a 30-day followup period following the initial *C. difficile* infection diagnosis. The Kaplan–Meier method was used to measure survival curves. In order to account for patients who exited the cohort during the followup period, patients were censored from the survival analysis following the last fact in their record. Use of de-identified, aggregate data was determined non-human subjects research (IRB-Non-HSR 22282) by the UVA Institutional Review Board for Health Sciences Research.

## Data availability

Data displayed and analyzed by the TriNetX Platform are in aggregate form or any patient-level data are de-identified due to protected health information. A detailed report of the TriNetX query and analysis (including propensity score density function plots before and after matching) are provided in the Supplementary Material. Contact R.P. for original data upon reasonable request.

## Results

94 healthcare organizations (100%) responded to the query with a total of 1,006,866 hospitalized *C. difficile* cases, 9,302 of which had anti-IL-23 medication (ustekinumab, guselkumab, or risankizumab) and 997,564 without anti-IL-23. Baseline characteristics, laboratory measurements, and comorbid conditions for the full cohort (before propensity matching) are shown in Table 1. 287/9,301 (3.1%) propensity-matched control *C. difficile* patients died within the 30 day followup period compared to 50/9,301 (0.54%) patients on at least one anti-IL-23 medication. The odds ratio for 30-day mortality in the anti-IL-23 group was 0.17 (95% confidence interval 0.126-0.230). A Kaplan Meier curve is shown in Figure 1, demonstrating significantly higher survival among anti-IL-23 treated patients (Log-rank *P*<0.001).

**Table 1.** Baseline characteristics of the full cohort.

|  | **Controls** | **Anti-IL-23** | **P value** |
| --- | --- | --- | --- |
| **A. Full Cohort** | n=996,414 | n=9,301 |  |
| **Demographics** |  |  |  |
| Age (mean+/-SD) | 58.7 +/- 20.8 | 43.2 +/- 18.5 | <0.001 |
| Female Sex | 526,595/996,414 (53.2) | 5,238/9,301 (56.3) | <0.001 |
| Race: Non-White | 108,946/925,909 (11.8) | 853/8,865 (9.6) | <0.001 |
| Ethnicity: Hispanic or Latino | 237,021/925,909 (25.6) | 1,581/8,865 (17.8) | <0.001 |
| **Comorbidities** |  |  |  |
| Inflammatory Bowel Disease (ICD-10 K50-K52) | 271,079/957,667 (28.3) | 8,412/9,076 (92.7) | <0.001 |
| Psoriasis (ICD-10 L40) | 18,739/957,667 (1.96) | 1,987/9,076 (21.9) | <0.001 |
| Diabetes (ICD-10 E08-E13) | 279,473/925,909 (30.2) | 1,711/8,865 (19.3) | <0.001 |
| Chronic Respiratory Disease (ICD-10 J40-J4A) | 271,657/925,909 (29.3) | 2,714/8,865 (30.6) | 0.008 |
| Cancer (ICD-10 C00-D49) | 394,073/925,909 (42.6) | 4,366/8,865 (49.2) | <0.001 |
| **Laboratory Measurements** | (Non-missing n) | (Non-missing n) |  |
| White Blood Cell Count (x10<sup>9</sup> cells/mL, mean+/-SD) | 13.4 +/- 42.3 (353,899) | 10.8 +/- 110 (8,416) | <0.001 |
| Creatinine (mg/dL, mean+/-SD) | 1.34 +/- 3.1 (743,106) | 1.16 +/- 5.8 (8,169) | <0.001 |
| Lactate (mmol/L, mean+/-SD) | 1.51 +/- 1.26 (340,938) | 1.29 +/- 0.674 (3,533) | <0.001 |
| Albumin (mg/dL, mean+/-SD) | 3.45 +/- 0.78 (715,511) | 3.8 +/- 0.65 (8,166) | <0.001 |
| **B. Propensity-Matched Cohort** | n=9,301 | n=9,301 |  |
| Age (mean+/-SD) | 43.2 +/- 18.5 | 43.2 +/- 18.5 | 1 |
| Female Sex | 5,238 (56.3) | 5,238 (56.3) | 1 |

Data shown as n/non-missing (%) unless otherwise specified.
Abbreviations: SD, standard deviation; International Classification of Diseases-10 code
(ICD-10).

**Figure 1.**
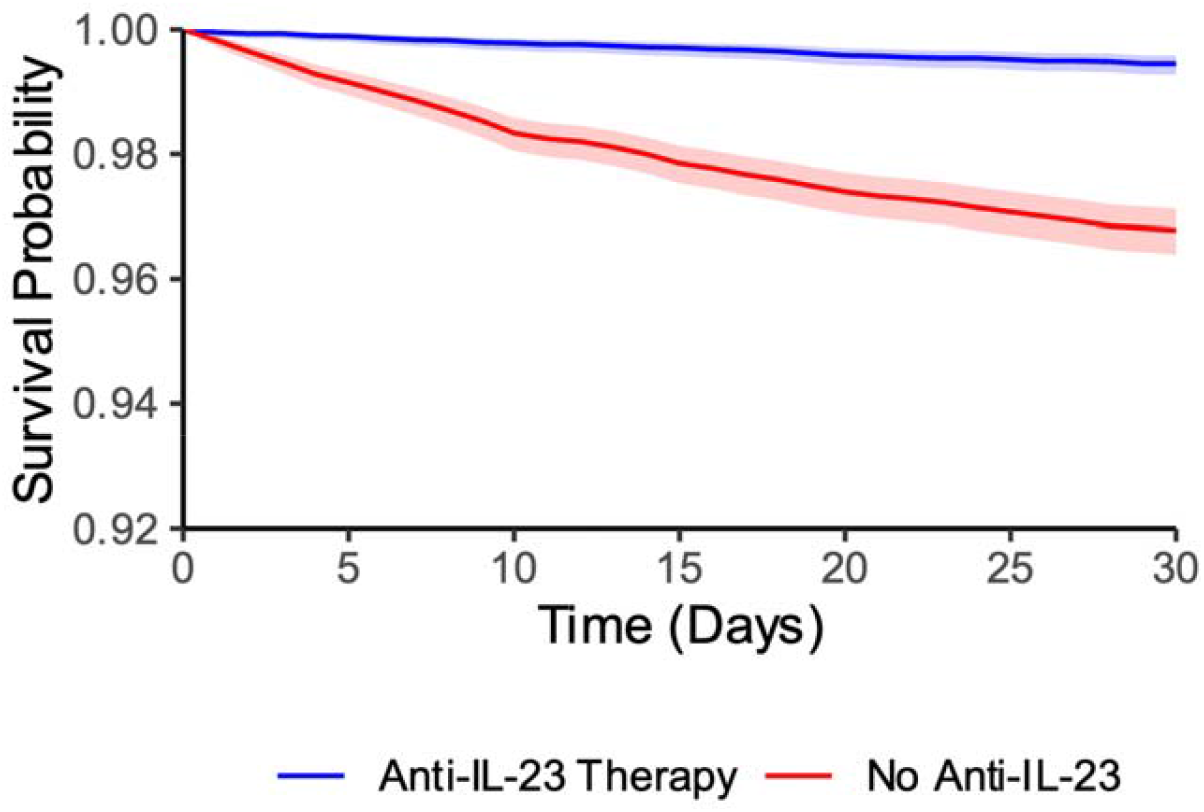
Kaplan Meier Survival Curve. Shaded areas represent the 95% confidence interval.

## Discussion

To our knowledge, this is the first observational study for the clinical use of host-directed immunotherapy in *C. difficile* infection suggesting that concomitant anti-IL-23 monoclonal antibodies may significantly reduce *C. difficile* infection mortality. Importantly, by enrolling patients on anti-IL-23 treatment strongly enriched those patients for its two major indications, inflammatory bowel disease (92.7%) and psoriasis (21.9%). *C. difficile* infection is not only more common with inflammatory bowel disease but also more deadly^10^ and psoriasis patients may be more likely to receive immunosuppressive medications that put them at higher risk for severe *C. difficile* infection. However, traditional baseline *C. difficile* severity markers^3^ (white blood cell count and creatinine) were both significantly lower in the anti-IL-23 group, suggesting that *C. difficile* on anti-IL-23 treatment may have been milder from the onset.

IL-23 inhibition offers several possible advantages and a better safety profile compared to conventional anti-*C. difficile* antibiotics, stool transplant, or surgery. Anti-IL-23 monoclonal antibodies have been in wide clinical use for over a decade and generally considered safe, with mostly benign side effects including nasopharyngitis, upper respiratory tract infection, and injection site erythema.^11^ Antibiotic exposure is the most important predisposing factor for *C. difficile* infection and anti-IL-23 monoclonal antibodies could spare patients from developing further antimicrobial resistance and microbiome disruption. Immunotherapy also does not risk introducing deadly nosocomial pathogens as from a stool transplant.^12^

This study has important limitations. As an observational study, there are likely sources of bias associated with anti-IL-23 treatment that were not captured or adjusted for in our analysis such as time-varying factors or a higher incidence of diabetes among non-anti-IL-23 patients. By virtue of selecting patients on anti-IL-23 therapy, the results may not be generalizable to other patient populations. In addition, identifying deaths directly attributable to *C. difficile* is notoriously difficult^13^ and therefore, we chose a relatively short (30-day) followup period in order to minimize deaths due to causes other than *C. difficile* infection. While billing codes are a reasonable proxy for identifying clinically relevant hospitalized *C. difficile* infection using large administrative databases such as TriNetX,^14^ some cases may have represented *C. difficile* colonization rather than infection.

While the TriNetX database offers one of the largest cohorts available to study outcomes of *C. difficile* infection, it has significant drawbacks including lack of direct access to patient-level identifiable information (including some characteristics of the matched control subcohort) and missing or heterogeneous data across organizations. Recurrent infection is a major issue with *C. difficile* infection but this outcome could not be meaningfully examined for two main reasons: the constraints of the TriNetX database in identifying time-varying endpoints besides mortality, and the poor accuracy of retrospective methods besides clinician chart review for identifying recurrence episodes.^15^

Anti-IL-23 is an attractive non-antibiotic, non-surgical candidate treatment for *C. difficile* infection based on our knowledge of *C. difficile* immunopathogenesis and over a decade of clinical experience with anti-IL-23 monoclonal antibodies. This retrospective study further supports the notion that IL-23 inhibition early in the course of *C. difficile* infection to prevent mortality is plausible and further human clinical trials are warranted.

## Disclosure of potential conflicts of interest

W. A. Petri is a consultant for TechLab Inc., a company that manufactures diagnostic tests for *C. difficile* toxins. All other authors report no conflicts of interest relevant to this article.

## Funding

This work was supported by the National Institutes of Health (K23AI163368 to G.R.M.; 5R01AI152477 and 5R01AI124214 to W.A.P).

## Author Contributions

G.R.M. and W.A.P. designed the study and G.R.M. wrote the manuscript. R.P. and S.P. conducted the TriNetX analysis. All authors contributed to the concepts, design, and analysis of the reported study.

## Notes

### Author Declarations

Use of de-identified, aggregate data was determined non-human subjects research (IRB-Non-HSR 22282) by the UVA Institutional Review Board for Health Sciences Research.

