## Supplementary Material for "Anti-Interleukin-23 Treatment Linked to Improved *Clostridioides difficile* Infection Survival"

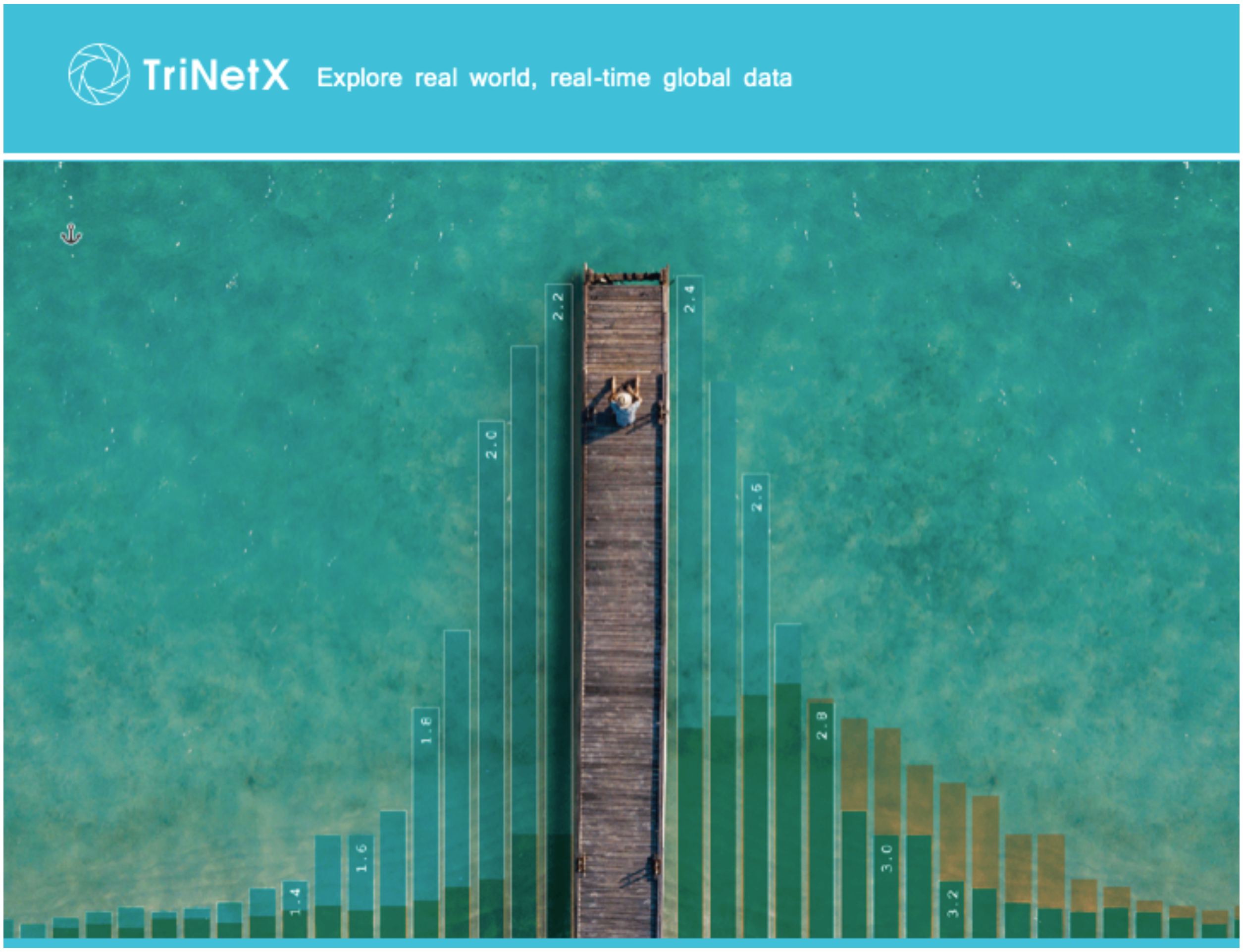


### various

#### Compare Outcomes Analysis

Created by TriNetX on Oct 9, 2024, 12:42:58 UTC

### Introduction

TriNetX is the global federated health research network providing access to electronic medical records (diagnoses, procedures, medications, laboratory values, genomic information) across large healthcare organizations (HCOs). This report was run on the set of HCOs grouped into a network called COVID-19 Research Network. This network included 94 HCO(s).

This report describes a Compare Outcomes Analysis, named IL23_difficile_inpatient_short, generated by the TriNetX platform on Oct 9, 2024, 12:42:58 UTC. This analysis compared the outcomes of two cohorts: Cohort A (9,302 patients) named +IL23_difficile_inpatient and Cohort B (997,564 patients) named -IL_difficile_inpatient.

This analysis was run by Robert Preissner and downloaded by Robert Preissner.

### Methods

The analysis process includes two main steps: 1) Defining the cohorts through query criteria; 2) Setting up and running the analysis. Setting up the analysis requires definitions for the index event, outcomes criteria, and the time frame. Compare outcomes supports four analyses: Measures of Association, Survival, Number of Instances and Lab result distribution. These analyses have additional options that are listed in the Outcomes Definitions and Analyses Specifications section below. Furthermore, characteristics of the cohorts that are balanced using propensity score matching are also included in the Propensity Score Matching section.

#### Cohorts definition

This section lists all terms used in the definitions of the two cohorts.

##### Query Criteria for Cohort 1 (query name: +IL23_difficile_inpatient)

This query was run on the network COVID-19 Research Network with 94 HCO(s) queried and 94 HCO(s) responded. A total of 59 provider(s) responded with patients. The final cohort included 9,302 patients who matched the query criteria listed in the table below. For the text representation of the query criteria please see Appendix A.

| **Ungrouped terms** | | | | |
|---|---|---|---|---|
| must have | any of | medication | NLM:RXNORM:847083 | ustekinumab |
| | | medication | NLM:RXNORM:1928588 | guselkumab |
| | | medication | NLM:RXNORM:2166040 | risankizumab |
| | and any of | procedure | UMLS:CPT:87493 | Infectious agent detection by nucleic acid (DNA or RNA); Clostridium difficile, toxin gene(s), amplified probe technique |
| | | laboratory | UMLS:LNC:54067-4 | Clostridioides difficile toxin genes [Presence] in Stool by NAA with probe detection |
| | | diagnosis | UMLS:ICD10CM:A04.7 | Enterocolitis due to Clostridium difficile |
| | | procedure | UMLS:CPT:87324 | Infectious agent antigen detection by immunoassay technique (eg, enzyme immunoassay [EIA], enzyme-linked |

| | | | | |
|---|---|---|---|---|
| | | | | immunosorbent assay [ELISA], fluorescence immunoassay [FIA], immunochemiluminometric assay [IMCA]), qualitative or semiquantitative; Clostridium difficile toxin(s) |
| | | diagnosis | UMLS:ICD10CM:A04.72 | Enterocolitis due to Clostridium difficile, not specified as recurrent |
| | | laboratory | UMLS:LNC:61367-9 | Clostridioides difficile DNA [Presence] in Specimen by NAA with probe detection |
| | and | visit | UMLS:HL7V3.0:VisitType:IMP | Visit: Inpatient Encounter |

##### Query Criteria for Cohort 2 (query name: -IL_difficile_inpatient)

This query was run on the network COVID-19 Research Network with 94 HCO(s) queried and 94 HCO(s) responded. A total of 81 provider(s) responded with patients. The final cohort included 997,564 patients who matched the query criteria listed in the table below.

| **Ungrouped terms** | | | | |
|---|---|---|---|---|
| must have | any of | procedure | UMLS:CPT:87493 | Infectious agent detection by nucleic acid (DNA or RNA); Clostridium difficile, toxin gene(s), amplified probe technique |
| | | laboratory | UMLS:LNC:54067-4 | Clostridioides difficile toxin genes [Presence] in Stool by NAA with probe detection |
| | | diagnosis | UMLS:ICD10CM:A04.7 | Enterocolitis due to Clostridium difficile |
| | | procedure | UMLS:CPT:87324 | Infectious agent antigen detection by immunoassay technique (eg, enzyme immunoassay [EIA], enzyme-linked immunosorbent assay [ELISA], fluorescence immunoassay [FIA], immunochemiluminometric assay [IMCA]), qualitative or semiquantitative; Clostridium difficile toxin(s) |
| | | diagnosis | UMLS:ICD10CM:A04.72 | Enterocolitis due to Clostridium difficile, not specified as recurrent |
| | | laboratory | UMLS:LNC:61367-9 | Clostridioides difficile DNA [Presence] in Specimen by NAA with probe detection |
| | and | visit | UMLS:HL7V3.0:VisitType:IMP | Visit: Inpatient Encounter |
| cannot have | | medication | NLM:RXNORM:847083 | ustekinumab |
| | or | medication | NLM:RXNORM:1928588 | guselkumab |
| | or | medication | NLM:RXNORM:2166040 | risankizumab |

#### Analysis Setup

This section contains the Index Event and Time Window definitions and a list of selected outcomes and the analyses.

##### Index Event & Time Window Definitions

The index event defines the point in time when each patient in the cohort enters the analysis. To define an index event for the cohort, one or more criteria for the cohort must be selected. The index date for each patient within a cohort is the day on which the patient first met the selected criteria for the cohort (listed in the table below).

As the index event defines the earliest time point after which outcomes are analyzed, the time window defines the duration during which outcomes are analyzed. The time window can start on the same day as the index event or at any specified time interval after the index event. The time window can end any time after the start date. Outcomes are defined as diagnoses, medications, procedures, or laboratory values that happened in the time window starting after the first occurrence of the index event.

##### Time Window Used in this Analysis

This analysis included outcomes that occurred in the time window that started 1 day after the first occurrence of the index event and ended 30 days after the first occurrence of the index event.

The index event only includes events that occurred up to 20 years ago. Patients whose index event occurred 20 years or more ago are excluded. In this analysis, 0 patients in Cohort 1 and 0 patients in Cohort 2 were excluded because they met the index event more than 20 years ago.

##### Index Events Used in this Analysis

Index events for the Compare Outcomes analysis were derived from the cohort definitions. Index events were defined separately for each cohort and were based on the criteria used in the original cohort definition. Please see Appendix B for the text representation of the index event definition.

The index event for Cohort 1 (query name: +IL23_difficile_inpatient) was defined as the following:

| **Ungrouped terms** | | | | |
|---|---|---|---|---|
| must have | any of | medication | NLM:RXNORM:847083 | ustekinumab |
| | | medication | NLM:RXNORM:1928588 | guselkumab |
| | | medication | NLM:RXNORM:2166040 | risankizumab |
| | and any of | procedure | UMLS:CPT:87493 | Infectious agent detection by nucleic acid (DNA or RNA); Clostridium difficile, toxin gene(s), amplified probe technique |
| | | laboratory | UMLS:LNC:54067-4 | Clostridioides difficile toxin genes [Presence] in Stool by NAA with probe detection |
| | | diagnosis | UMLS:ICD10CM:A04.7 | Enterocolitis due to Clostridium difficile |
| | | procedure | UMLS:CPT:87324 | Infectious agent antigen detection by immunoassay technique (eg, enzyme immunoassay [EIA], enzyme-linked immunosorbent assay [ELISA], fluorescence immunoassay [FIA], immunochemiluminometric assay [IMCA]), qualitative or semiquantitative; Clostridium difficile toxin(s) |
| | | diagnosis | UMLS:ICD10CM:A04.72 | Enterocolitis due to Clostridium difficile, not specified as recurrent |
| | | laboratory | UMLS:LNC:61367-9 | Clostridioides difficile DNA [Presence] in Specimen by NAA with probe detection |
| | and | visit | UMLS:HL7V3.0:VisitType:IMP | Visit: Inpatient Encounter |

The index event for Cohort 2 (query name: -IL_difficile_inpatient) was defined as the following:

| **Ungrouped terms** | | | | |
|---|---|---|---|---|
| must have | any of | procedure | UMLS:CPT:87493 | Infectious agent detection by nucleic acid (DNA or RNA); Clostridium difficile, |

| | | | |
|---|---|---|---|
| | | | toxin gene(s), amplified probe technique |
| | laboratory | UMLS:LNC:54067-4 | Clostridioides difficile toxin genes [Presence] in Stool by NAA with probe detection |
| | diagnosis | UMLS:ICD10CM:A04.7 | Enterocolitis due to Clostridium difficile |
| | procedure | UMLS:CPT:87324 | Infectious agent antigen detection by immunoassay technique (eg, enzyme immunoassay [EIA], enzyme-linked immunosorbent assay [ELISA], fluorescence immunoassay [FIA], immunochemiluminometric assay [IMCA]), qualitative or semiquantitative; Clostridium difficile toxin(s) |
| | diagnosis | UMLS:ICD10CM:A04.72 | Enterocolitis due to Clostridium difficile, not specified as recurrent |
| | laboratory | UMLS:LNC:61367-9 | Clostridioides difficile DNA [Presence] in Specimen by NAA with probe detection |
| and | visit | UMLS:HL7V3.0:VisitType:IMP | Visit: Inpatient Encounter |

#### Analyses Specifications

The Compare Outcomes Analytic supports four types of analyses: Measure of Association, Survival, Number of Instances, and Lab result distribution. The first three analyses support the “exclude patients with outcomes prior to the window” setting. This option can exclude patients from the analysis if they are not at risk for an outcome (e.g., if the outcome is a chronic disease). When "exclude patients with the outcome prior to the time window" is not checked, all patients in the cohort are included in the analysis, regardless of whether they had the outcome prior to the time window. When "exclude patients with the outcome prior to the time window" is checked, patients are excluded from the analysis if their record includes the outcome prior to the beginning of the time window. This selection will exclude all patients with the outcome prior to the index event. If the start of the time window for the analysis falls some days after the index event, patients will also be excluded if they have the outcome between the index event and the start of the time window.

#### Measure of Association Analysis

The Measure of Association Analysis calculates and compares the fraction of patients with the selected outcome. The output summary includes: Patients in each Cohort (count of patients meeting query criteria); Patients with Outcome in each Cohort (of the patients in the cohort, count of patients that had the outcome in the time window); and Risk (the fraction of patients in the cohort that have the outcome in the time window, i.e. Patients with Outcome / Patients in Cohort). In addition, Risk Difference (the difference in the risks in Cohort 1 and Cohort 2), Risk Ratio (the ratio of the risks in Cohort 1 and Cohort 2), and Odds Ratio (the ratio of the odds in Cohort 1 and Cohort 2). The bar chart shows the risk of the outcome for the both cohorts.

#### Survival Analysis

The Kaplan-Meier Analysis estimates probability of the outcome at a respective time interval (daily time interval is used in this analysis). In order to account for the patients who exited the cohort during the analysis period, and therefore should not be included in the analysis, censoring is applied. In this analysis, patients are removed from the analysis (censored) after the last fact in their record.

The output summary includes: Patients in each Cohort (count of patients meeting query criteria); Patients with Outcome (of the patients in the cohort, count of patients that had the outcome in the time window); Median Survival (the number of days when the survival drops below 50%; the “-” indicates that survival does not drop below 50% during the time window); and Survival Probability at End of Time Window (the % survival at the end of the time window). In addition, Log-Rank test, Hazard Ratio and test for Proportionality.

##### Number of Instances Analysis

The Number of Instances Analysis calculates how many times the outcome occurred in the time window. This analysis includes two additional settings: include patients with zero instances; the definition of an instance.

Selecting to exclude patients with zero instances will remove these patients from the calculations for mean number of instances, standard deviation, or median. The histogram showing the distribution of patients by number of instances will not contain a bar for zero. Alternatively, by selecting to include patients with zero instances, the mean, standard deviation, and median for number of instances will reflect these patients. The histogram will contain a bar for zero patients.

The definition of an instance affects how counts are analyzed. By selecting Date, each calendar date on which any of the terms selected in the outcome are recorded will represent one instance. For example, if the outcome is “Med A or Med B,” and a patient has “Med A” on January 3, then both medications on January 4, then “Med B” on January 6, then that patient is considered to have three instances– January 3, January 4, and January 6. Note that if an outcome occurs across several dates (e.g. Visit: inpatient encounter), then only the start date is tracked for the purpose of counting instances. A patient who begins at stay on January 1, ends that stay on January 3, begins another stay on January 10, and ends that stay on January 15, is considered to have two instances of the outcome.

Selecting Visit as an Instance will count any visit that includes the outcome as one instance, regardless of how many times it occurred. For instance, consider a patient administered an analgesic on each of the three days that make up an inpatient stay following some index event. If analgesic is an outcome, these three administrations will represent only one instance, because all three are associated with the same visit.

The output summary includes: Patients in Cohort (count of patients meeting query criteria); Patients with Outcome (of the patients in the cohort, count of patients that had the outcome in the time window); Mean (mean of the counts); Standard Deviation (standard deviation of the counts); Median (median of the counts); and Median (1+ instances) when patients with zero instances included in the analysis. In addition, T-Test statistics testing for the difference between the cohorts is included.

##### Laboratory Results Analysis

Lab Results can be included in the analysis only for the outcomes that are labs. Only the most recent lab values in the time window are included. For the lab results that are numeric, the outcome summary includes: Patients in Cohort (count of patients meeting query criteria); Patients with Outcome (of the patients in the cohort, count of patients that had the outcome in the time window); Mean (mean of the counts); and Standard Deviation (the standard deviation for lab values across patients in the cohort). In addition, T-Test statistics testing for the difference between the cohorts is included.

For the non-numeric lab results, three values are reported: counts of Negative; Positives; and Unknowns. The counts are represented in the bar chart as percentages of the total counts.

#### Outcome Definitions

Table below outlines the definitions for each outcome and the analysis specifications. For outcome definitions consisting of more than one term, at least one term must match. Please see Appendix C for the text representation of the outcome definitions.

| death | | |
|---|---|---|
| **Outcome definition** | | |
| Demographics | Deceased | Deceased |
| **Settings for the performed analyses** | | |
| Risk analysis | | including patients with outcome prior to the time window |
| Kaplan - Meier survival analysis | | including patients with outcome prior to the time window |

#### Propensity Score Matching

Propensity score matching was performed on all listed characteristics. Characteristics of the cohorts before and after matching are summarized in the table below.

**Cohort 1 and cohort 2 patient count before and after propensity score matching**

| Cohort | Patient count before matching | Patient count after matching |
|---|---|---|
| 1 - +IL23_difficile_inpatient | 9,301 | 9,301 |
| 2 - -IL_difficile_inpatient | 996,414 | 9,301 |

**Propensity score density function - Before and after matching (cohort 1 - purple, cohort 2 - green)**

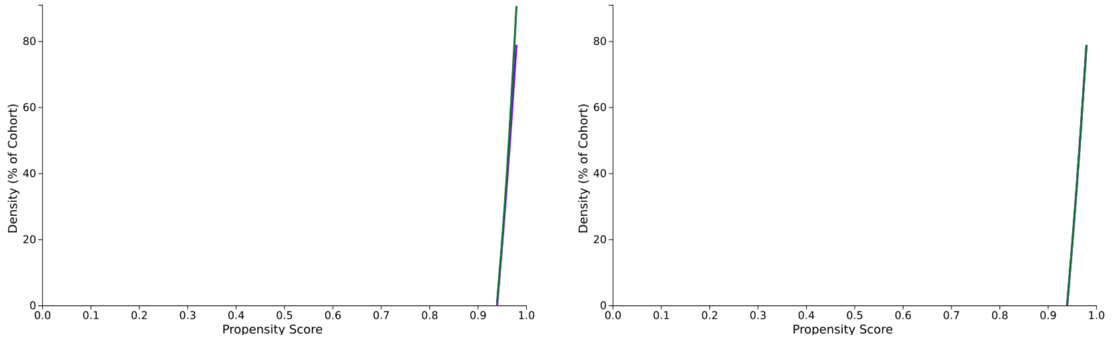


**Cohort 1 (N = 9,301) and cohort 2 (N = 996,414) characteristics before propensity score matching**

**Demographics**

| Cohort | | | Mean ± SD | Patients | % of Cohort | P-Value | Std diff. |
|---|---|---|---|---|---|---|---|
| 1<br>2 | AI | Age at Index | 43.2 +/- 18.5<br>58.7 +/- 20.8 | 9,301<br>989,452 | 100%<br>100% | <0.001 | 0.788 |
| 1<br>2 | F | Female | | 5,238<br>526,595 | 56.3%<br>53.2% | <0.001 | 0.062 |

**Cohort 1 (N = 9,301) and cohort 2 (N = 9,301) characteristics after propensity score matching**

**Demographics**

| Cohort | | | Mean ± SD | Patients | % of Cohort | P-Value | Std diff. |
|---|---|---|---|---|---|---|---|
| 1<br>2 | AI | Age at Index | 43.2 +/- 18.5<br>43.2 +/- 18.5 | 9,301<br>9,301 | 100%<br>100% | 1 | <0.001 |
| 1<br>2 | F | Female | | 5,238<br>5,238 | 56.3%<br>56.3% | 1 | <0.001 |

### Results

Results are summarized in the tables below.  Outcomes analysis was performed on the cohorts after propensity score matching.

#### 1 death

##### Risk analysis

| Cohort | | Patients in cohort | Patients with outcome | Risk |
|---|---|---|---|---|
| 1 | +IL23_difficile_inpatient | 9,301 | 50 | 0.005 |
| 2 | -IL_difficile_inpatient | 9,301 | 287 | 0.031 |

| | | 95% CI | z | p |
|---|---|---|---|---|
| **Risk Difference** | -0.025 | (-0.029, -0.022) | -13.029 | 0.000 |
| **Risk Ratio** | 0.174 | (0.129, 0.235) | N/A | N/A |
| **Odds Ratio** | 0.170 | (0.126, 0.230) | N/A | N/A |

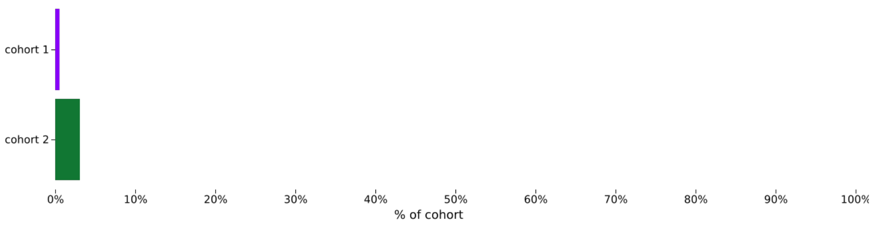


##### Kaplan - Meier survival analysis

| Cohort | | Patients in cohort | Patients with outcome | Median survival (days) | Survival probability at end of time window |
|---|---|---|---|---|---|
| 1 | +IL23_difficile_inpatient | 9,301 | 50 | -- | 99.45% |
| 2 | -IL_difficile_inpatient | 9,301 | 287 | -- | 96.78% |

| | $\chi^2$ | df | p |
|---|---|---|---|
| **Log-Rank Test** | 176.380 | 1 | 0.000 |

| | Hazard Ratio | 95% CI | $\chi^2$ | df | p |
|---|---|---|---|---|---|
| **Hazard Ratio and Proportionality** | 0.167 | (0.124, 0.226) | 1.887 | 1 | 0.170 |

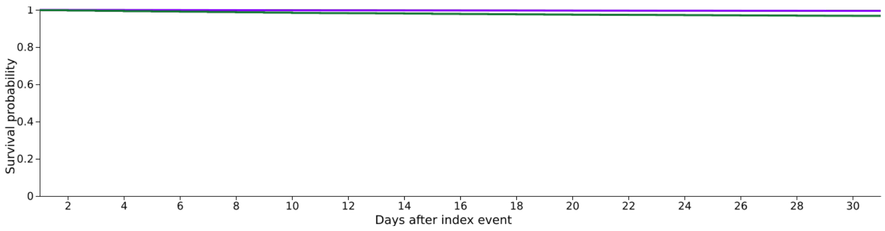
Survival probability
1
0.8
0.6
0.4
0.2
0
2
4
6
8
10
12
14
16
18
20
22
24
26
28
30
Days after index event

### Appendix A – Text Representation of the Cohorts Definition

This section lists all terms used in the definitions of the two cohorts.

#### Query Criteria for Cohort 1 (query name: +IL23_difficile_inpatient)

Patients must have:
all of the following:
any of the following:
ustekinumab (NLM:RXNORM:847083); or
guselkumab (NLM:RXNORM:1928588); or
risankizumab (NLM:RXNORM:2166040); and
any of the following:
Infectious agent detection by nucleic acid (DNA or RNA); Clostridium difficile, toxin gene(s), amplified probe technique (UMLS:CPT:87493); or
Clostridioides difficile toxin genes [Presence] in Stool by NAA with probe detection (UMLS:LNC:54067-4); or
Enterocolitis due to Clostridium difficile (UMLS:ICD10CM:A04.7); or
Infectious agent antigen detection by immunoassay technique (eg, enzyme immunoassay [EIA], enzyme-linked immunosorbent assay [ELISA], fluorescence immunoassay [FIA], immunochemiluminometric assay [IMCA]), qualitative or semiquantitative; Clostridium difficile toxin(s) (UMLS:CPT:87324); or
Enterocolitis due to Clostridium difficile, not specified as recurrent (UMLS:ICD10CM:A04.72); or
Clostridioides difficile DNA [Presence] in Specimen by NAA with probe detection (UMLS:LNC:61367-9); and
Visit: Inpatient Encounter (UMLS:HL7V3.0:VisitType:IMP).

#### Query Criteria for Cohort 2 (query name: -IL_difficile_inpatient)

Patients must have:
all of the following:
any of the following:
Infectious agent detection by nucleic acid (DNA or RNA); Clostridium difficile, toxin gene(s), amplified probe technique (UMLS:CPT:87493); or
Clostridioides difficile toxin genes [Presence] in Stool by NAA with probe detection (UMLS:LNC:54067-4); or
Enterocolitis due to Clostridium difficile (UMLS:ICD10CM:A04.7); or
Infectious agent antigen detection by immunoassay technique (eg, enzyme immunoassay [EIA], enzyme-linked immunosorbent assay [ELISA], fluorescence immunoassay [FIA], immunochemiluminometric assay [IMCA]), qualitative or semiquantitative; Clostridium difficile toxin(s) (UMLS:CPT:87324); or
Enterocolitis due to Clostridium difficile, not specified as recurrent (UMLS:ICD10CM:A04.72); or
Clostridioides difficile DNA [Presence] in Specimen by NAA with probe detection (UMLS:LNC:61367-9); and
Visit: Inpatient Encounter (UMLS:HL7V3.0:VisitType:IMP).

Patients cannot have:
any of the following:
ustekinumab (NLM:RXNORM:847083); or
guselkumab (NLM:RXNORM:1928588); or
risankizumab (NLM:RXNORM:2166040).

### Appendix B – Text Representation of the Analysis Setup

This section contains the Index Event definition for each cohort.

The index event for Cohort 1 (query name: +IL23_difficile_inpatient) is defined as the following:

Patients must have:
  all of the following:
    any of the following:
      ustekinumab (NLM:RXNORM:847083); or
      guselkumab (NLM:RXNORM:1928588); or
      risankizumab (NLM:RXNORM:2166040); and
    any of the following:
      Infectious agent detection by nucleic acid (DNA or RNA); Clostridium difficile, toxin gene(s), amplified probe technique (UMLS:CPT:87493); or
      Clostridioides difficile toxin genes [Presence] in Stool by NAA with probe detection (UMLS:LNC:54067-4); or
      Enterocolitis due to Clostridium difficile (UMLS:ICD10CM:A04.7); or
      Infectious agent antigen detection by immunoassay technique (eg, enzyme immunoassay [EIA], enzyme-linked immunosorbent assay [ELISA], fluorescence immunoassay [FIA], immunochemiluminometric assay [IMCA]), qualitative or semiquantitative; Clostridium difficile toxin(s) (UMLS:CPT:87324); or
      Enterocolitis due to Clostridium difficile, not specified as recurrent (UMLS:ICD10CM:A04.72); or
      Clostridioides difficile DNA [Presence] in Specimen by NAA with probe detection (UMLS:LNC:61367-9); and
    Visit: Inpatient Encounter (UMLS:HL7V3.0:VisitType:IMP).

The index event for Cohort 2 (query name: -IL_difficile_inpatient) is defined as the following:

Patients must have:
  all of the following:
    any of the following:
      Infectious agent detection by nucleic acid (DNA or RNA); Clostridium difficile, toxin gene(s), amplified probe technique (UMLS:CPT:87493); or
      Clostridioides difficile toxin genes [Presence] in Stool by NAA with probe detection (UMLS:LNC:54067-4); or
      Enterocolitis due to Clostridium difficile (UMLS:ICD10CM:A04.7); or
      Infectious agent antigen detection by immunoassay technique (eg, enzyme immunoassay [EIA], enzyme-linked immunosorbent assay [ELISA], fluorescence immunoassay [FIA], immunochemiluminometric assay [IMCA]), qualitative or semiquantitative; Clostridium difficile toxin(s) (UMLS:CPT:87324); or
      Enterocolitis due to Clostridium difficile, not specified as recurrent (UMLS:ICD10CM:A04.72); or
      Clostridioides difficile DNA [Presence] in Specimen by NAA with probe detection (UMLS:LNC:61367-9); and
    Visit: Inpatient Encounter (UMLS:HL7V3.0:VisitType:IMP).

### Appendix C – Text Representation of the Outcomes Definition

This analysis includes the following outcomes:

death

Patients must have:
  Deceased (Deceased).
